# Rapid base-specific calling of SARS-CoV-2 variants of concern using combined RT-PCR melting curve screening and SIRPH technology

**DOI:** 10.1101/2021.03.15.21253586

**Authors:** Tierling Sascha, Kattler Kathrin, Vogelgesang Markus, Pfuhl Thorsten, Lohse Stefan, Lo Porto Christina, Schmitt Beate, Salhab Abdulrahman, Smola Sigrun, Walter Jörn

## Abstract

The emergence of novel variants of concern of SARS-CoV-2 demands a fast and reliable detection of such variants in local populations. Here we present a cost-efficient and fast workflow combining a pre-screening of SARS-CoV-2 positive samples using RT-PCR melting curve analysis with multiplexed IP-RP-HPLC-based single nucleotide primer extensions (SIRPH). The entire workflow from positive SARS-CoV-2 testing to base-specific identification of variants requires about 24 h. We applied the sensitive method to monitor the local VOC outbreaks in a few hundred positive samples collected in a confined region of Germany.

## Background

During the ongoing COVID-19 pandemic recent reports of new SARS-CoV-2 variants with changes in viral functionalities or pathogenic aspects have caused major concern. In December 2020 several emerging variants of concern (VOCs) with increased transmission were reported from UK (B.1.1.7), South Africa (B.1.351) and Brazil (P.1). To allow the timely adaptation of public health interventions it is of utmost importance to monitor and precisely follow the ingression and distribution of these VOCs as quickly as possible. They are each characterized by a set of mutations that can be used for variant monitoring. Apart from the spike-protein mutation N501Y in all three currently circulating VOCs, B.1.351 and P.1 also share the mutations K417N/T and E484K, but are discriminated by the mutation V1176F only present in P.1 [1-3]. The amino acid substitution E484K in the receptor-binding domain of the spike-protein was reported to be associated with immune escape from neutralizing antibodies which is especially worrisome regarding vaccination or reinfection [4].

The gold standard for the identification of virus variants is sequencing of full viral genomes. Currently, RT-PCR melting curve analysis approaches followed by confirmation through viral genome sequencing are widely used. However, the required time for NGS sequencing and the extra costs ask for faster and more cost-effective test systems. Previously, we proposed a pooling approach for high-throughput RT-PCR testing to identify SARS-CoV-2 infection in large cohorts of asymptomatic people [5]. Here we extend this strategy to obtain a fast and accurate base-specific detection of VOCs by combining a sequential RT-PCR based melting curve analysis as a pre-screening approach of SARS-CoV-2 positive samples with a multiplexed IP-RP-HPLC-based single nucleotide primer extension approach (SIRPH, 6, 7]. This protocol allows a fast and comprehensive generation of “single base sequencing” signatures for a relevant spectrum of VOCs to assign the variant lineages within the (local) pandemic situation.

## METHODS

### SARS-CoV-2 (pool)-testing and N501Y RT-PCR melting curve analysis

Respiratory samples were tested individually (expected high positivity rate) or in pools (expected low positivity rate, 16 samples per pool) performed with Biomek i5 Span8 (Beckman Coulter) by automated SARS-CoV-2 dual target RT-PCR (E and ORF1) using Cobas 6800 (Roche Diagnostics). To identify VOC candidates positive samples were extracted using RNAdvance Viral and Biomek i5 MC96 (Beckman Coulter) and subjected to N501Y RT-PCR melting curve analysis (VirSNiP SARS-CoV-2 Spike N501Y, TibMolBiol) using LightCycler® 480 Instrument II (Roche Life Sciences). All analyses were performed according to the manufacturers’ instructions. The study was approved by the local ethics committee of the Saarland University Medical Center at Saarland Ärztekammer.

### Reverse Transcription

Isolated RNA was reverse transcribed using random hexamer primers (NEB) and Maxima Reverse Transcriptase H- (ThermoFisher) following the manufacturer’s recommendations. cDNA from the same preparation was submitted to PCRs for SNuPE and NGS Library generation.

### SNuPE and IP-RP-HPLC (SIRPH)

Two µl of cDNA were used as template in a 30 μl reaction in the presence of 3 mM Tris–HCl (pH 8.8), 0.7 mM (NH4)2SO4, 50 mM KCl, 2.5 mM MgCl2, 0.06 mM of each dNTP, 3 U HotFire DNA polymerase (Solis BioDyne) and 167 nM primers (Sup. Table 1). PCRs were performed at 95° C for 15 min followed by 35 cycles 95°C/60 s, 54°C (58°C for A570D+D614G) /30 s, 72°C/30 s and a final extension 72°C/5 min. Five μl of PCR products were treated with 1U of ExoCIAP (mixture of Exonuclease I (Jena Bioscience) and Calf Alkaline Intestine Phosphatase (Calbiochem)) for 30 min at 37°C. To inactivate the ExoCIP enzymes the reaction was incubated for 15 min at 80°C. Afterwards, 14 μl primer extension mastermix (50 mM Tris–HCl, pH 9.5, 2.5 mM MgCl2, 0.05 mM of ddNTPs, 1.6 μM of each SNuPE primer, 2.5 U Termipol DNA polymerase (Solis BioDyne) was added. Primer extension reactions were performed at 96°C for 2 min followed by 50 cycles 96°C/30 s, 50°C/30 s, 60°C/20 s. Separation of products was conducted on a XBridge BEH C18 2.5µm 4.6mmx50mm column (Waters) at 0.9 ml/min and 50°C by continuously mixing buffer B (0.1 M TEAA, 25% acetonitril) to buffer A (0.1 M TEAA, Sup. Table 2). Mutations were determined by relative retention times in comparison to the non-mutated reference.

### Sanger sequencing

Sanger sequencing of exemplary PCR products generated for SIRPH analysis was performed at Macrogen (Amsterdam) using 40 ng purified PCR product and 10 pmol of corresponding PCR primers.

### Viral genome sequencing

NGS Libraries for viral genome sequencing were generated using the primer design from the ARTIC v0.3 protocol [8] modified for Illumina sequencing. We added Illumina compatible adapter sequences directly to the ARTIC Primers (Sup. Table 3). To obtain a more equal coverage we introduced a third multiplex pool with 5 primer pairs covering difficult genomic regions (Sup. Table 3). Multiplex PCRs were carried out as described in the ARTIC protocol. After purification PCR products were submitted to an indexing PCR with TruSeq primers (Illumina) for 8 cycles. After final purification and normalization samples were sequenced on a MiSeq (Illumina).

### NGS data processing

Viral genome data were processed using CoVpipe (https://gitlab.com/RKIBioinformaticsPipelines/ncov_minipipe). After trimming of adapter/primer sequences and low quality reads (<Q30), reads were aligned to the SARS-CoV-2 reference NC_045512.2. Reads mapping to the human genome were excluded. Variant calling was performed with default parameter and consensus sequences were masked for lowly covered sites (<20X). Visualization was achieved with Auspice and Nextstrain [9].

## RESULTS

Single nucleotide primer extension (SNuPE) coupled to IP-RP-HPLC detection (SIRPH) is a powerful and fast approach for point mutation analysis of known genomic variants. This assay is characterized by primers that anneal with their 3’-end directly adjacent to potentially mutated sites (PMS). Using ddNTPs the primer can only be extended by a single nucleotide complementary to the genomic information on the annealed DNA strand. Due to the incorporation of different bases in wildtype and mutant, extended primers differ in hydrophobicity which allows their separation by IP-RP-HPLC. We adapted this approach for screening of VOCs in the SARS-CoV-2 genome in a multiplex assay that allows simultaneous detection of several mutations.

We established the method on 80 SARS-CoV-2 positive samples collected in the south-west of Germany (Saarland) from Nov/Dec 2020 for B.1.1.7, B.1.351 and P.1 (Figure 1A). We designed primers for our SNuPE assay (Figure 1B/C, Sup. Table 1) to determine the nucleotide at the PMS D614G (A→G in all clades except 19A), N501Y present in all 3 currently circulating VOCs (A→T) and the discriminatory PMS A570D (C→A in B.1.1.7), P681H (C→A in B.1.1.7), T716I (C→T in B.1.1.7), E484K (G→A in B.1.351 and P.1) as well as V11176T (G→T in P.1). Besides, we analyzed two local mutations, i.e. L9P (T→C) and G172R (G→C) that we observed in a high frequency in NGS data of earlier samples (Sep/Oct 2020, Figure 1D) from Saarland to follow their further ingression and distribution.

**Figure 1.**
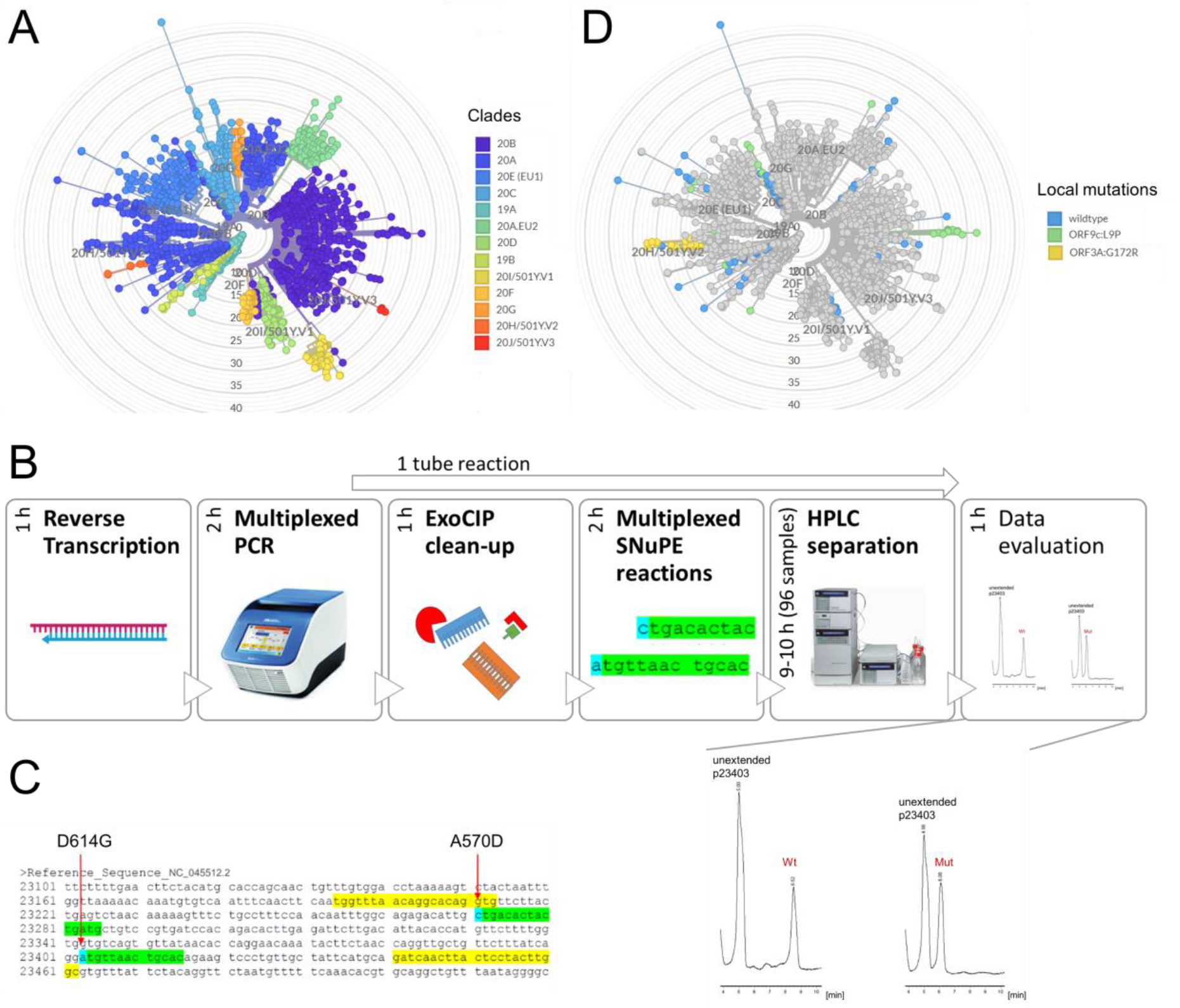
Multiplexed IP-RP-HPLC-based single nucleotide primer extension approach (SIRPH). A – Phylogenetic tree of SARS-CoV-2 sequences derived from GISAID references (accessed on 2021/01/29) and sequencing data from Saarland (March – October 2020, available on GISAID using identifiers listed in Sup. Table 6) colored by clade. Variants of concern are found in clades 20I/501Y.V1 (B.1.1.7), 20H/501Y.V2 (B.1.351) and 20J/501Y.V3 (P.1). B – SIRPH Workflow: Isolated RNA is reverse transcribed, followed by a multiplexed PCR of regions of interest. After ExoCIP clean-up of residual dNTPs and primers, multiplexed SNuPE reactions are performed that are separated by IP-RP-HPLC. The whole workflow can be performed within 1 day. C – Exemplary SNuPE primer design (green) for the PMS (blue) A570D and D614G within a single PCR product (yellow). D – Distribution of the local mutations ORF9c:L9P (green) and ORF3A:G172R (yellow) in samples from Saarland (March – October 2020). E – Exemplary HPLC separation of SNuPE reactions at the PMS D614G showing a profile for wildtype (Wt, A) and mutation (Mut, G).

Notably, mutations in close vicinity of each other that could be covered within the same PCR product (Figure 1C, Sup. Figure 1) were assessed simultaneously in a multiplexed SNuPE reaction. After RNA extraction and reverse transcription, cDNA was used as template for four different PCR reactions yielding PCR products harbouring the PMS (Figure 1B). In a following one-tube reaction PCR products were treated with exonuclease I and calf intestinal phosphatase (ExoCIP) to remove primers and nucleotides, heat-inactivated and subjected to SNuPE followed by IP-RP-HPLC separation (Figure 1B) in 96 sample formats. In these SNuPE reactions we either assessed two PMS at once (E484K, N501Y, P681H, T716I, L9P, G172R) or one PMS (A570D, D614G) per reaction (Sup. Figure 1) resulting in a total of five HPLC runs per sample.

All samples showed the mutation D614G which emerged early in the pandemic and is now predominant in samples from across the globe [10]. The PMS characteristic for the three analyzed VOCs (E484K, N501Y, P681H, T716I, A570D) were in concordance with the original SARS-CoV-2 sequence (Sup. Table 4). Our local mutations revealed interesting patterns over time (Sup. Table 4). The spread of the ORF9c mutation L9P dropped from 35.3% in Sep/Oct 2020 (n = 51) to 6.25 % in Nov/Dec 2020 (n = 80) while the ORF3A mutation G172R remained more abundant (33.3% to 27.5%).

For validation we performed full viral genome sequencing on all 80 samples. For variant calling we only considered sites with coverage of at least 20 (Sup. Table 4). Especially in the region harbouring our local L9P variant we observed high coverage-related drop-out rates in the sequencing data. Thus, we performed additional validation of exemplary samples by Sanger sequencing (Sup. Figure 2). All SNuPE results were in full concordance with viral genome sequencing and Sanger sequencing-based variant calling (Sup. Table 4, Sup. Figure 2) demonstrating the reliability and robustness of our SNuPE approach.

Inspired by these results we developed a rapid screening strategy for prospective high-throughput SARS-CoV-2 and subsequent VOC testing (Figure 2A). Asymptomatic individuals, i.e. health care workers of the local University Hospital, were screened by SARS-CoV-2 pool testing in pools of 16, while samples from symptomatic people or public health services (contact tracing) with expected positivity rates above 2% were investigated by individual SARS-CoV-2 RT-PCR. Positive nucleic acid extracts were immediately subjected to N501Y mutation specific RT-PCR melting curve analysis to identify candidates of currently circulating VOCs. Subsequent SNuPE analysis was performed to clearly identify and assign the respective VOC.

**Figure 2.**
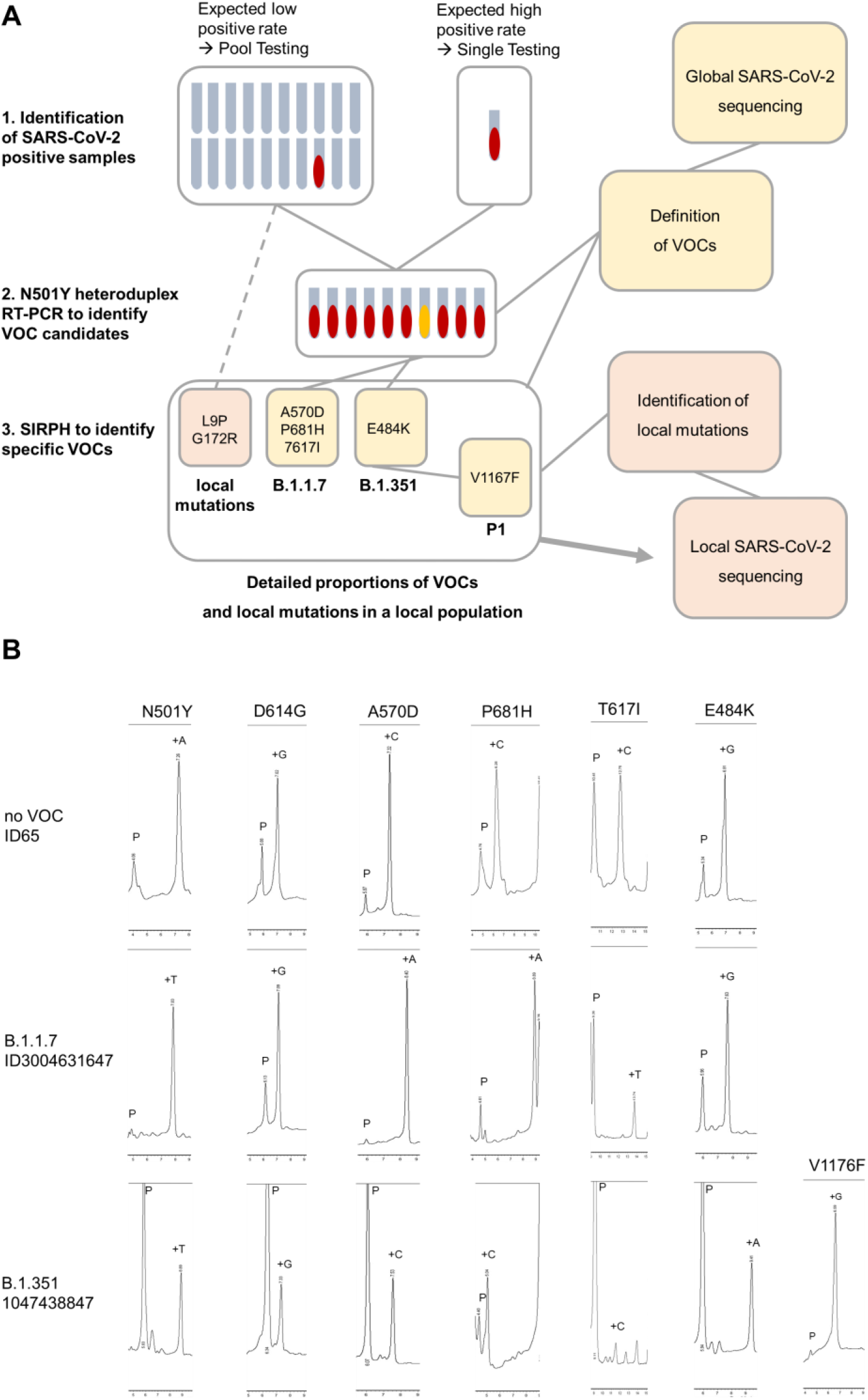
High-throughput screening strategy for variant monitoring in a local population. A – Iterative Workflow from Identification of SARS-CoV-2 positive samples (1) followed by variant candidate identification by RT-PCR melting curve analysis (2) and final identification of specific VOCs as defined by global sequencing efforts. Detailed proportions of VOCs or local variants of interest identified by SIRPH analysis can then be used for selection of samples in a local sequencing projects. B – HPLC separation profiles of the VOC characterizing PMS set for an exemplary non-VOC (ID65), a B.1.1.7 and a B.1.351 variant as identified in the workflow described in A.

We applied this approach for a routine test on a large cohort to identify B.1.1.7, B.1.351 and P.1 variants in Saarland generating reliable and highly accurate results. From 2059 pooled and 2987 individual respiratory samples collected between January, 25th and 31st 2021, 542 samples were tested SARS-CoV-2 positive by RT-PCR. Within these we identified 13 VOC candidates by RT-PCR melting curve analysis based screening during this early phase of VOC introduction into Saarland. All candidates were submitted to SIRPH analysis for confirmation and further VOC assignment revealing the presence of all characteristic mutations 501Y, 570D, 681H and 716I in 12 out of 13 pre-defined samples clearly classifying them as B.1.1.7, while 1 sample was identified as B.1.351 (Figure 2B, Sup. Table 6). The whole process lasted about 24 hours from positive RT-PCR testing to PMS sequence validation.

## DISCUSSION

Our presented strategy shows that a combination of pre-screening by (pooled) SARS-CoV-2 and subsequent N501Y RT-PCR melting curve analysis together with a confirmatory SIRPH analysis generates fast and reliable results to speed up monitoring of VOCs in larger cohorts. We demonstrate how this strategy can quickly be implemented for VOC monitoring to rationalize local public health interventions by supporting the tracing of VOCs. While whole-genome sequencing will remain the gold standard for virus monitoring and the detection of new mutations, the proposed screening by PCR/SIRPH has several advantages. First of all the method is robust, fast (24h) and very cost-effective compared to NGS confirmation. Notably, it robustly calls regions with coverage drop-outs in sequencing data (Sup. Figure 2). It can even be applied to samples with low viral load (Ct ≤ 33) that typically fail or result in low quality data during sequencing (Ct > 27). Furthermore the method is flexible and can be implemented on a variety of existing RP-HPLC systems. In addition, we perform SNuPE assays as “one-tube” reactions that can be multiplexed. New mutation specific assays can be rapidly established within two working days to include newly emerging variants allowing a fast and precise population surveillance. The combined pool PCR/SIRPH approach can be used to flank sequencing surveillance programs where usually only up to 5% of randomly chosen positive cases will be full-genome sequenced. The emergence and spreading of local variants asks for such fast and deep monitoring in the entire population in a timely manner. Since time is crucial for surveillance, our method offers base-specific results in one day as compared to NGS-based sequencing which usually require up to 5 working days. Ongoing sequencing of full viral genomes in a proportion of samples will be used to screen for the development of new SARS-CoV-2 variants. In an iterative process the information gained by sequencing can be used to easily adapt the SIRPH approach to new VOCs that may emerge in the future course of the pandemic.

## Data Availability

Fasta files of full viral genome sequencing can be accessed on GISAID (https://www.gisaid.org/) using the identifiers listed in Sup. Table 6.

## ACKNOWLEDGEMENT

The authors wish to acknowledge the contributions of the technicians Katrin Thieser, Luisa Scheidhauer, Helga Appel and Barbara Best.

## Footnotes

### Mention of any meeting(s) where the information has previously been presented

Not applicable.

