## Supplement for "Rapid base-specific calling of SARS-CoV-2 variants of concern using combined RT-PCR melting curve screening and SIRPH technology"

**Supplementary Figure 1**: SIRPH Primer design for PCR (yellow) and SNuPE (green). Potentially mutated sites are marked in blue. Primer sequences are listed in Sup. Table 1.

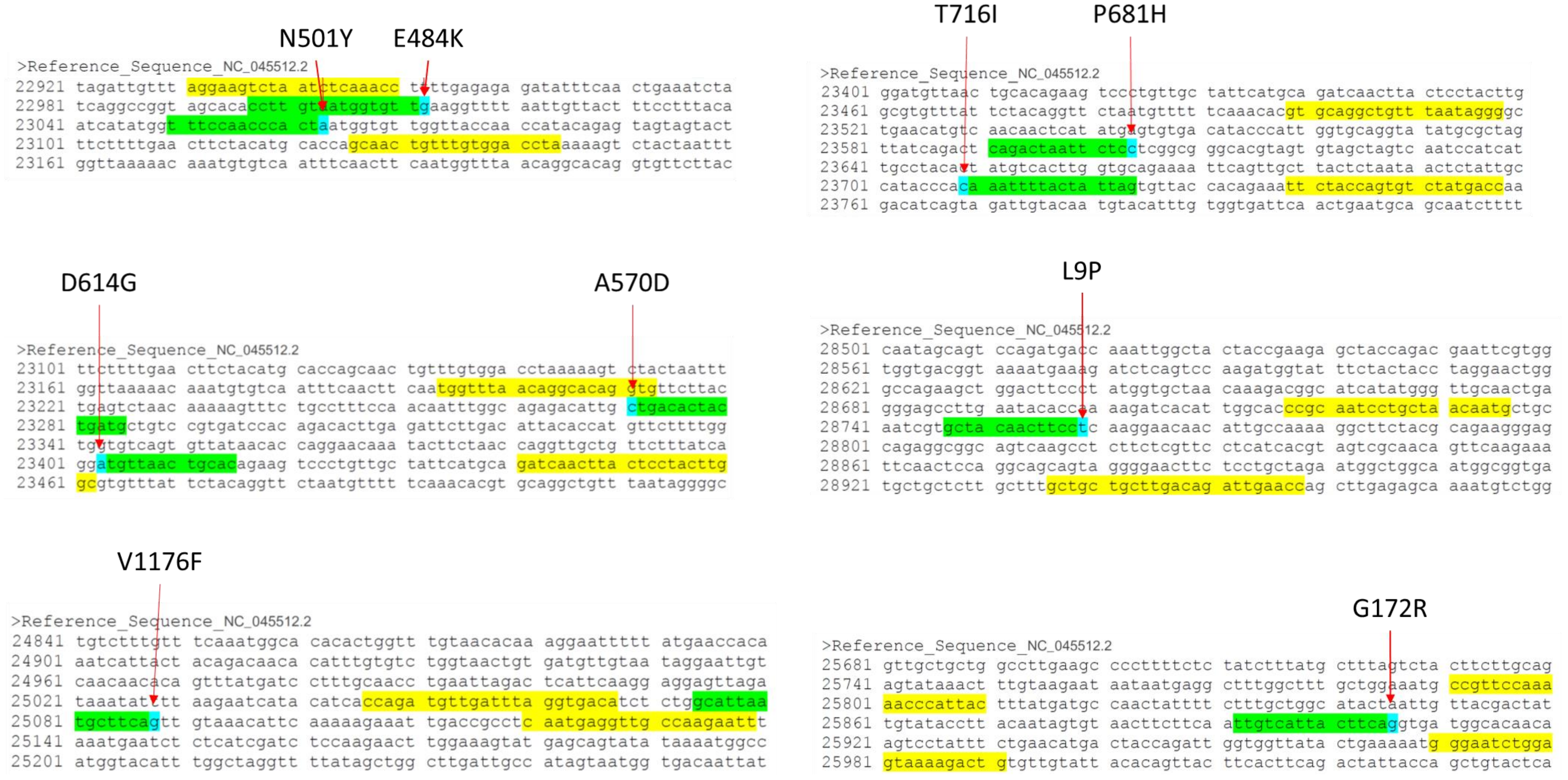

**Supplementary Figure 2:** Exemplary results of Sanger sequencing for potentially mutated sites L9P (marked in blue).

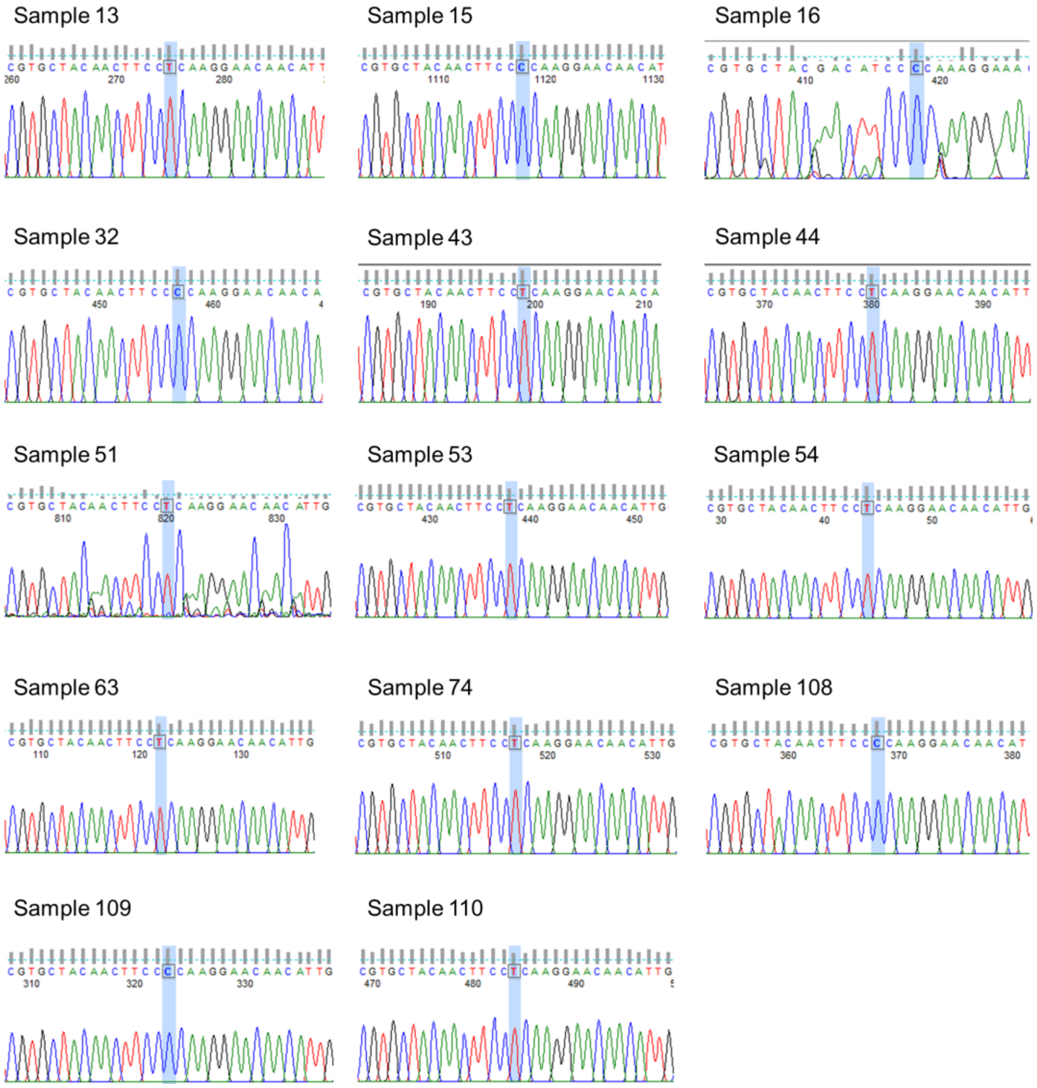

**Supplementary Table 1:** Primers for mutation screening via PCR and SNuPE. SNuPE primers are printed in bold.

| | |
|---|---|
| N501Y+E484K F | 5´-AGGAAGTCTAATCTCAAACC-3´ |
| N501Y+E484K R | 5´-TAGGTCCACAAACAGTTGC-3´ |
| **E484K nt23012** | **5´-CCTTGTAATGGTGTT-3´** |
| **N501Y nt23063** | **5´-TTTCCAACCCACT-3´** |
| A570D+D614G F | 5´-TGGTTTAACAGGCACAGGTG-3´ |
| A570D+D614G R | 5´-GCCAAGTAGGAGTAAGTTGATC-3´ |
| **A570D nt23271** | **5´-CATCAGTAGTGTCA-3´** |
| **D614G nt23403** | **5´-GTGCAGTTAACA-3´** |
| P681H+T716I F | 5´-GTGCAGGCTGTTTAATAGGG-3´ |
| P681H+T716I R | 5´-GGTCATAGACACTGGTAGAA-3´ |
| **P681H nt23604** | **5´-CAGACTAATTCTC-3´** |
| **T716I nt23709** | **5´-CTAATAGTAAAATTT-3´** |
| L9P F | 5´-CCGCAATCCTGCTAACAATG-3´ |
| L9P R | 5´-GGTTCAATCTGTCAAGCAGCAGC-3´ |
| **L9P nt28759** | **5´-GCTACAACTTCC-3´** |
| G172R F | 5´-CCGTTCCAAAAACCCATTAC-3´ |
| G172R R | 5´-CAGTCTTTTACTCCAGATTCCC-3´ |
| **G172R nt25906** | **5´-TTGTCATTACTTCA-3´** |

**Supplementary Table 2:** Acetonitril gradients for HPLC separation of SNuPE products.

| | |
|---|---|
| N501Y+E484K | 29-35% buffer B over 10 min |
| A570D | 27-34% buffer B over 8 min |
| D614G | 27-32% buffer B over 8 min |
| P681H+T716I | 28-35% buffer B over 13 min |
| L9P+G172R | 27-38% buffer B over 12 min |

**Supplementary Table 3:** Artic multiplex PCR primers for SARS-CoV-2 genome sequencing adapted for Illumina. Low coverage regions were amplified in a separate pool (Pool 3) to obtain a more equal sequencing depth across the genome.

| | | |
|---|---|---|
| COVID19_1_F | TCTTTCCCTACACGACGCTCTTCCGATCTACCAACCAACTTTCGATCTCTTGT | Pool 1 |
| COVID19_1_R | GTGACTGGAGTTCAGACGTGTGCTCTTCCGATCTCATCTTTAAGATGTTGACGTGCCTC | Pool 1 |
| COVID19_2_F | TCTTTCCCTACACGACGCTCTTCCGATCTCTGTTTTACAGGTTCGCGACGT | Pool 2 |
| COVID19_2_R | GTGACTGGAGTTCAGACGTGTGCTCTTCCGATCTTAAGGATCAGTGCCAAGCTCGT | Pool 2 |
| COVID19_3_F | TCTTTCCCTACACGACGCTCTTCCGATCTCGGTAATAAAGGAGCTGGTGGC | Pool 1 |
| COVID19_3_R | GTGACTGGAGTTCAGACGTGTGCTCTTCCGATCTAAGGTGTCTGCAATTCATAGCTCT | Pool 1 |
| COVID19_4_F | TCTTTCCCTACACGACGCTCTTCCGATCTGGTGTATACTGCTGCCGTGAAC | Pool 2 |
| COVID19_4_R | GTGACTGGAGTTCAGACGTGTGCTCTTCCGATCTCACAAGTAGTGGCACCTTCTTTAGT | Pool 2 |
| COVID19_5_F | TCTTTCCCTACACGACGCTCTTCCGATCTTGGTGAAACTTCATGGCAGACG | Pool 1 |

| | | |
|---|---|---|
| COVID19_5_R | GTGACTGGAGTTCAGACGTGTGCTCTTCCGATCTATTGATGTTGACTTTCTCTTTTTGGAGT | Pool 1 |
| COVID19_6_F | TCTTTCCCTACACGACGCTCTTCCGATCTGGTGTTGTTGGAGAAGGTTCCG | Pool 2 |
| COVID19_6_R | GTGACTGGAGTTCAGACGTGTGCTCTTCCGATCTTAGCGGCCTTCTGTAAAACACG | Pool 2 |
| COVID19_7_F | TCTTTCCCTACACGACGCTCTTCCGATCTATCAGAGGCTGCTCGTGTTGTA | Pool 1 |
| COVID19_7_F_alt0 | TCTTTCCCTACACGACGCTCTTCCGATCTCATTTGCATCAGAGGCTGCTCG | Pool 1 |
| COVID19_7_R | GTGACTGGAGTTCAGACGTGTGCTCTTCCGATCTTGCACAGGTGACAATTTGTCCA | Pool 1 |
| COVID19_7_R_alt5 | GTGACTGGAGTTCAGACGTGTGCTCTTCCGATCTAGGTGACAATTTGTCCACCGAC | Pool 1 |
| COVID19_8_F | TCTTTCCCTACACGACGCTCTTCCGATCTAGAGTTTCTTAGAGACGGTTGGGA | Pool 3 |
| COVID19_8_R | GTGACTGGAGTTCAGACGTGTGCTCTTCCGATCTGCTTCAACAGCTTCACTAGTAGGT | Pool 3 |
| COVID19_9_F | TCTTTCCCTACACGACGCTCTTCCGATCTTCCCACAGAAGTGTTAACAGAGGA | Pool 1 |
| COVID19_9_F_alt4 | TCTTTCCCTACACGACGCTCTTCCGATCTTTCCCACAGAAGTGTTAACAGAGG | Pool 1 |
| COVID19_9_R | GTGACTGGAGTTCAGACGTGTGCTCTTCCGATCTATGACAGCATCTGCCACAACAC | Pool 1 |
| COVID19_9_R_alt2 | GTGACTGGAGTTCAGACGTGTGCTCTTCCGATCTGACAGCATCTGCCACAACACACAG | Pool 1 |
| COVID19_10_F | TCTTTCCCTACACGACGCTCTTCCGATCTTGAGAAGTGCTCTGCCTATACAGT | Pool 2 |
| COVID19_10_R | GTGACTGGAGTTCAGACGTGTGCTCTTCCGATCTTCATCTAACCAATCTTCTTCTTGCTCT | Pool 2 |
| COVID19_11_F | TCTTTCCCTACACGACGCTCTTCCGATCTGGAATTTGGTGCCACTTCTGCT | Pool 1 |
| COVID19_11_R | GTGACTGGAGTTCAGACGTGTGCTCTTCCGATCTTCATCAGATTCAACTTGCATGGCA | Pool 1 |
| COVID19_12_F | TCTTTCCCTACACGACGCTCTTCCGATCTAAACATGGAGGAGGTGTTGCAG | Pool 2 |
| COVID19_12_R | GTGACTGGAGTTCAGACGTGTGCTCTTCCGATCTTTCACTCTTCATTTCCAAAAAGCTTGA | Pool 2 |
| COVID19_13_F | TCTTTCCCTACACGACGCTCTTCCGATCTTCGCACAAATGTCTACTTAGCTGT | Pool 1 |
| COVID19_13_R | GTGACTGGAGTTCAGACGTGTGCTCTTCCGATCTACCACAGCAGTTAAAACACCCT | Pool 1 |

| | | |
|---|---|---|
| COVID19_14_F | TCTTTCCCTACACGACGCTCTTCCGATCTCATCCAGATTCTGCCACTCTTGT | Pool 2 |
| COVID19_14_F_alt4 | TCTTTCCCTACACGACGCTCTTCCGATCTTGGCAATCTTCATCCAGATTCTGC | Pool 2 |
| COVID19_14_R | GTGACTGGAGTTCAGACGTGTGCTCTTCCGATCTAGTTTCCACACAGACAGGCATT | Pool 2 |
| COVID19_14_R_alt2 | GTGACTGGAGTTCAGACGTGTGCTCTTCCGATCTTGCGTGTTTCTTCTGCATGTGC | Pool 2 |
| COVID19_15_F | TCTTTCCCTACACGACGCTCTTCCGATCTACAGTGCTTAAAAAGTGTAAAAGTGCC | Pool 1 |
| COVID19_15_F_alt1 | TCTTTCCCTACACGACGCTCTTCCGATCTAGTGCTTAAAAAGTGTAAAAGTGCCT | Pool 1 |
| COVID19_15_R | GTGACTGGAGTTCAGACGTGTGCTCTTCCGATCTAACAGAAACTGTAGCTGGCACT | Pool 1 |
| COVID19_15_R_alt3 | GTGACTGGAGTTCAGACGTGTGCTCTTCCGATCTACTGTAGCTGGCACTTTGAGAGA | Pool 1 |
| COVID19_16_F | TCTTTCCCTACACGACGCTCTTCCGATCTAATTTGGAAGAAGCTGCTCGGT | Pool 2 |
| COVID19_16_R | GTGACTGGAGTTCAGACGTGTGCTCTTCCGATCTCACAACTTGCGTGTGGAGGTTA | Pool 2 |
| COVID19_17_F | TCTTTCCCTACACGACGCTCTTCCGATCTCTTCTTTCTTTGAGAGAAGTGAGGACT | Pool 1 |
| COVID19_17_R | GTGACTGGAGTTCAGACGTGTGCTCTTCCGATCTTTTGTTGGAGTGTTAACAATGCAGT | Pool 1 |
| COVID19_18_F | TCTTTCCCTACACGACGCTCTTCCGATCTTGGAAATACCCACAAGTTAATGGTTTAAC | Pool 2 |
| COVID19_18_F_alt2 | TCTTTCCCTACACGACGCTCTTCCGATCTACTTCTATTAAATGGGCAGATAACAACTGT | Pool 2 |
| COVID19_18_R | GTGACTGGAGTTCAGACGTGTGCTCTTCCGATCTAGCTTGTTTACCACACGTACAAGG | Pool 2 |
| COVID19_18_R_alt1 | GTGACTGGAGTTCAGACGTGTGCTCTTCCGATCTGCTTGTTTACCACACGTACAAGG | Pool 2 |
| COVID19_19_F | TCTTTCCCTACACGACGCTCTTCCGATCTGCTGTTATGTACATGGGCACACT | Pool 1 |
| COVID19_19_R | GTGACTGGAGTTCAGACGTGTGCTCTTCCGATCTTGTCCAACTTAGGGTCAATTTCTGT | Pool 1 |
| COVID19_20_F | TCTTTCCCTACACGACGCTCTTCCGATCTACAAAGAAAACAGTTACACAACAACCA | Pool 2 |
| COVID19_20_R | GTGACTGGAGTTCAGACGTGTGCTCTTCCGATCTACGTGGCTTTATTAGTTGCATTGTT | Pool 2 |
| COVID19_21_F | TCTTTCCCTACACGACGCTCTTCCGATCTTGGCTATTGATTATAAACACTACACACCC | Pool 1 |

| | | |
|---|---|---|
| COVID19_21_F_alt2 | TCTTTCCCTACACGACGCTCTTCCGATCTGGCTATTGATTATAAACACTACACACCCT | Pool 1 |
| COVID19_21_R | GTGACTGGAGTTCAGACGTGTGCTCTTCCGATCTTAGATCTGTGTGGCCAACCTCT | Pool 1 |
| COVID19_21_R_alt0 | GTGACTGGAGTTCAGACGTGTGCTCTTCCGATCTGATCTGTGTGGCCAACCTCTTC | Pool 1 |
| COVID19_22_F | TCTTTCCCTACACGACGCTCTTCCGATCTACTACCGAAGTTGTAGGAGACATTATACT | Pool 2 |
| COVID19_22_R | GTGACTGGAGTTCAGACGTGTGCTCTTCCGATCTACAGTATTCTTTGCTATAGTAGTCGGC | Pool 2 |
| COVID19_23_F | TCTTTCCCTACACGACGCTCTTCCGATCTACAACTACTAACATAGTTACACGGTGT | Pool 1 |
| COVID19_23_R | GTGACTGGAGTTCAGACGTGTGCTCTTCCGATCTACCAGTACAGTAGGTTGCAATAGTG | Pool 1 |
| COVID19_24_F | TCTTTCCCTACACGACGCTCTTCCGATCTAGGCATGCCTTCTTACTGTACTG | Pool 2 |
| COVID19_24_R | GTGACTGGAGTTCAGACGTGTGCTCTTCCGATCTACATTCTAACCATAGCTGAAATCGGG | Pool 2 |
| COVID19_25_F | TCTTTCCCTACACGACGCTCTTCCGATCTGCAATTGTTTTTCAGCTATTTTGCAGT | Pool 1 |
| COVID19_25_R | GTGACTGGAGTTCAGACGTGTGCTCTTCCGATCTACTGTAGTGACAAGTCTCTCGCA | Pool 1 |
| COVID19_26_F | TCTTTCCCTACACGACGCTCTTCCGATCTTTGTGATACATTCTGTGCTGGTAGT | Pool 2 |
| COVID19_26_R | GTGACTGGAGTTCAGACGTGTGCTCTTCCGATCTTCCGCACTATCACCAACATCAG | Pool 2 |
| COVID19_27_F | TCTTTCCCTACACGACGCTCTTCCGATCTACTACAGTCAGCTTATGTGTCAACC | Pool 1 |
| COVID19_27_R | GTGACTGGAGTTCAGACGTGTGCTCTTCCGATCTAATACAAGCACCAAGGTCACGG | Pool 1 |
| COVID19_28_F | TCTTTCCCTACACGACGCTCTTCCGATCTACATAGAAGTTACTGGCGATAGTTGT | Pool 2 |
| COVID19_28_R | GTGACTGGAGTTCAGACGTGTGCTCTTCCGATCTTGTTTAGACATGACATGAACAGGTGT | Pool 2 |
| COVID19_29_F | TCTTTCCCTACACGACGCTCTTCCGATCTACTTGTGTTCCTTTTTGTTGCTGC | Pool 1 |
| COVID19_29_R | GTGACTGGAGTTCAGACGTGTGCTCTTCCGATCTAGTGTACTCTATAAGTTTTGATGGTGTGT | Pool 1 |
| COVID19_30_F | TCTTTCCCTACACGACGCTCTTCCGATCTGCACAACTAATGGTGACTTTTTGCA | Pool 2 |
| COVID19_30_R | GTGACTGGAGTTCAGACGTGTGCTCTTCCGATCTACCACTAGTAGATACACAAACACCAG | Pool 2 |

| | | |
|---|---|---|
| COVID19_31_F | TCTTTCCCTACACGACGCTCTTCCGATCTTTCTGAGTACTGTAGGCACGGC | Pool 1 |
| COVID19_31_R | GTGACTGGAGTTCAGACGTGTGCTCTTCCGATCTACAGAATAAACACCAGGTAAGAATGAGT | Pool 1 |
| COVID19_32_F | TCTTTCCCTACACGACGCTCTTCCGATCTTGGTGAATACAGTCATGTAGTTGCC | Pool 2 |
| COVID19_32_R | GTGACTGGAGTTCAGACGTGTGCTCTTCCGATCTAGCACATCACTACGCAACTTTAGA | Pool 2 |
| COVID19_33_F | TCTTTCCCTACACGACGCTCTTCCGATCTACTTTTGAAGAAGCTGCGCTGT | Pool 1 |
| COVID19_33_R | GTGACTGGAGTTCAGACGTGTGCTCTTCCGATCTTGGACAGTAAACTACGTCATCAAGC | Pool 1 |
| COVID19_34_F | TCTTTCCCTACACGACGCTCTTCCGATCTTCCCATCTGGTAAAGTTGAGGGT | Pool 2 |
| COVID19_34_R | GTGACTGGAGTTCAGACGTGTGCTCTTCCGATCTAGTGAAATTGGGCCTCATAGCA | Pool 2 |
| COVID19_35_F | TCTTTCCCTACACGACGCTCTTCCGATCTTGTTCGCATTCAACCAGGACAG | Pool 1 |
| COVID19_35_R | GTGACTGGAGTTCAGACGTGTGCTCTTCCGATCTACTTCATAGCCACAAGGTTAAAGTCA | Pool 1 |
| COVID19_36_F | TCTTTCCCTACACGACGCTCTTCCGATCTTTAGCTTGGTTGTACGCTGCTG | Pool 2 |
| COVID19_36_R | GTGACTGGAGTTCAGACGTGTGCTCTTCCGATCTGAACAAAGACCATTGAGTACTCTGGA | Pool 2 |
| COVID19_37_F | TCTTTCCCTACACGACGCTCTTCCGATCTACACACCACTGGTTGTTACTCAC | Pool 1 |
| COVID19_37_R | GTGACTGGAGTTCAGACGTGTGCTCTTCCGATCTGTCCACACTCTCCTAGCACCAT | Pool 1 |
| COVID19_38_F | TCTTTCCCTACACGACGCTCTTCCGATCTACTGTGTTATGTATGCATCAGCTGT | Pool 2 |
| COVID19_38_R | GTGACTGGAGTTCAGACGTGTGCTCTTCCGATCTCACCAAGAGTCAGTCTAAAGTAGCG | Pool 2 |
| COVID19_39_F | TCTTTCCCTACACGACGCTCTTCCGATCTAGTATTGCCCTATTTTCTTCATAACTGGT | Pool 1 |
| COVID19_39_R | GTGACTGGAGTTCAGACGTGTGCTCTTCCGATCTTGTAACTGGACACATTGAGCCC | Pool 1 |
| COVID19_40_F | TCTTTCCCTACACGACGCTCTTCCGATCTTGCACATCAGTAGTCTTACTCTCAGT | Pool 2 |
| COVID19_40_R | GTGACTGGAGTTCAGACGTGTGCTCTTCCGATCTCATGGCTGCATCACGGTCAAAT | Pool 2 |
| COVID19_41_F | TCTTTCCCTACACGACGCTCTTCCGATCTGTTCCCTTCCATCATATGCAGCT | Pool 1 |

| | | |
|---|---|---|
| COVID19_41_R | GTGACTGGAGTTCAGACGTGTGCTCTTCCGATCTTGGTATGACAACCATTAGTTTGGCT | Pool 1 |
| COVID19_42_F | TCTTTCCCTACACGACGCTCTTCCGATCTTGCAAGAGATGGTTGTGTTCCC | Pool 2 |
| COVID19_42_R | GTGACTGGAGTTCAGACGTGTGCTCTTCCGATCTCCTACCTCCCTTTGTTGTGTTGT | Pool 2 |
| COVID19_43_F | TCTTTCCCTACACGACGCTCTTCCGATCTTACGACAGATGTCTTGTGCTGC | Pool 1 |
| COVID19_43_R | GTGACTGGAGTTCAGACGTGTGCTCTTCCGATCTAGCAGCATCTACAGCAAAAGCA | Pool 1 |
| COVID19_44_F | TCTTTCCCTACACGACGCTCTTCCGATCTTGCCACAGTACGTCTACAAGCT | Pool 2 |
| COVID19_44_F_alt3 | TCTTTCCCTACACGACGCTCTTCCGATCTCCACAGTACGTCTACAAGCTGG | Pool 2 |
| COVID19_44_R | GTGACTGGAGTTCAGACGTGTGCTCTTCCGATCTAACCTTTCCACATACCGCAGAC | Pool 2 |
| COVID19_44_R_alt0 | GTGACTGGAGTTCAGACGTGTGCTCTTCCGATCTCGCAGACGGTACAGACTGTGTT | Pool 2 |
| COVID19_45_F | TCTTTCCCTACACGACGCTCTTCCGATCTTACCTACAACTTGTGCTAATGACCC | Pool 1 |
| COVID19_45_F_alt2 | TCTTTCCCTACACGACGCTCTTCCGATCTAGTATGTACAAATACCTACAACTTGTGCT | Pool 1 |
| COVID19_45_R | GTGACTGGAGTTCAGACGTGTGCTCTTCCGATCTAAATTGTTTCTTCATGTTGGTAGTTAGAGA | Pool 1 |
| COVID19_45_R_alt7 | GTGACTGGAGTTCAGACGTGTGCTCTTCCGATCTTTCATGTTGGTAGTTAGAGAAAGTGTGTC | Pool 1 |
| COVID19_46_F | TCTTTCCCTACACGACGCTCTTCCGATCTTGTCGCTTCCAAGAAAAGGACG | Pool 2 |
| COVID19_46_F_alt1 | TCTTTCCCTACACGACGCTCTTCCGATCTCGCTTCCAAGAAAAGGACGAAGA | Pool 2 |
| COVID19_46_R | GTGACTGGAGTTCAGACGTGTGCTCTTCCGATCTCACGTTCACCTAAGTTGGCGTA | Pool 2 |
| COVID19_46_R_alt2 | GTGACTGGAGTTCAGACGTGTGCTCTTCCGATCTCACGTTCACCTAAGTTGGCGTAT | Pool 2 |
| COVID19_47_F | TCTTTCCCTACACGACGCTCTTCCGATCTAGGACTGGTATGATTTTGTAGAAAACCC | Pool 1 |
| COVID19_47_R | GTGACTGGAGTTCAGACGTGTGCTCTTCCGATCTAATAACGGTCAAAGAGTTTTAACCTCTC | Pool 1 |
| COVID19_48_F | TCTTTCCCTACACGACGCTCTTCCGATCTTGTTGACACTGACTTAACAAAGCCT | Pool 2 |
| COVID19_48_R | GTGACTGGAGTTCAGACGTGTGCTCTTCCGATCTTAGATTACCAGAAGCAGCGTGC | Pool 2 |

| | | |
|---|---|---|
| COVID19_49_F | TCTTTCCCTACACGACGCTCTTCCGATCTAGGAATTACTTGTGTATGCTGCTGA | Pool 1 |
| COVID19_49_R | GTGACTGGAGTTCAGACGTGTGCTCTTCCGATCTTGACGATGACTTGGTTAGCATTAATACA | Pool 1 |
| COVID19_50_F | TCTTTCCCTACACGACGCTCTTCCGATCTGTTGATAAGTACTTTGATTGTTACGATGGT | Pool 2 |
| COVID19_50_R | GTGACTGGAGTTCAGACGTGTGCTCTTCCGATCTTAACATGTTGTGCCAACCACCA | Pool 2 |
| COVID19_51_F | TCTTTCCCTACACGACGCTCTTCCGATCTTCAATAGCCGCCACTAGAGGAG | Pool 1 |
| COVID19_51_R | GTGACTGGAGTTCAGACGTGTGCTCTTCCGATCTAGTGCATTAACATTGGCCGTGA | Pool 1 |
| COVID19_52_F | TCTTTCCCTACACGACGCTCTTCCGATCTCATCAGGAGATGCCACAACTGC | Pool 2 |
| COVID19_52_R | GTGACTGGAGTTCAGACGTGTGCTCTTCCGATCTGTTGAGAGCAAAATTCATGAGGTCC | Pool 2 |
| COVID19_53_F | TCTTTCCCTACACGACGCTCTTCCGATCTAGCAAAATGTTGGACTGAGACTGA | Pool 1 |
| COVID19_53_R | GTGACTGGAGTTCAGACGTGTGCTCTTCCGATCTAGCCTCATAAAACTCAGGTTCCC | Pool 1 |
| COVID19_54_F | TCTTTCCCTACACGACGCTCTTCCGATCTTGAGTTAACAGGACACATGTTAGACA | Pool 2 |
| COVID19_54_R | GTGACTGGAGTTCAGACGTGTGCTCTTCCGATCTAACCAAAAACTTGTCCATTAGCACA | Pool 2 |
| COVID19_55_F | TCTTTCCCTACACGACGCTCTTCCGATCTACTCAACTTTACTTAGGAGGTATGAGCT | Pool 1 |
| COVID19_55_R | GTGACTGGAGTTCAGACGTGTGCTCTTCCGATCTGGTGTACTCTCCTATTTGTACTTTACTGT | Pool 1 |
| COVID19_56_F | TCTTTCCCTACACGACGCTCTTCCGATCTACCTAGACCACCACTTAACCGA | Pool 2 |
| COVID19_56_R | GTGACTGGAGTTCAGACGTGTGCTCTTCCGATCTACACTATGCGAGCAGAAGGGTA | Pool 2 |
| COVID19_57_F | TCTTTCCCTACACGACGCTCTTCCGATCTATTCTACACTCCAGGGACCACC | Pool 1 |
| COVID19_57_R | GTGACTGGAGTTCAGACGTGTGCTCTTCCGATCTGTAATTGAGCAGGGTCGCCAAT | Pool 1 |
| COVID19_58_F | TCTTTCCCTACACGACGCTCTTCCGATCTTGATTTGAGTGTTGTCAATGCCAGA | Pool 2 |
| COVID19_58_R | GTGACTGGAGTTCAGACGTGTGCTCTTCCGATCTCTTTTCTCCAAGCAGGGTTACGT | Pool 2 |
| COVID19_59_F | TCTTTCCCTACACGACGCTCTTCCGATCTTCACGCATGATGTTTCATCTGCA | Pool 1 |

| | | |
|---|---|---|
| COVID19_59_R | GTGACTGGAGTTCAGACGTGTGCTCTTCCGATCTAAGAGTCCTGTTACATTTTCAGCTTG | Pool 1 |
| COVID19_60_F | TCTTTCCCTACACGACGCTCTTCCGATCTTGATAGAGACCTTTATGACAAGTTGCA | Pool 2 |
| COVID19_60_R | GTGACTGGAGTTCAGACGTGTGCTCTTCCGATCTGGTACCAACAGCTTCTCTAGTAGC | Pool 2 |
| COVID19_61_F | TCTTTCCCTACACGACGCTCTTCCGATCTTGTTTATCACCCGCGAAGAAGC | Pool 1 |
| COVID19_61_R | GTGACTGGAGTTCAGACGTGTGCTCTTCCGATCTATCACATAGACAACAGGTGCGC | Pool 1 |
| COVID19_62_F | TCTTTCCCTACACGACGCTCTTCCGATCTGGCACATGGCTTTGAGTTGACA | Pool 2 |
| COVID19_62_R | GTGACTGGAGTTCAGACGTGTGCTCTTCCGATCTGTTGAACCTTTCTACAAGCCGC | Pool 2 |
| COVID19_63_F | TCTTTCCCTACACGACGCTCTTCCGATCTTGTTAAGCGTGTTGACTGGACT | Pool 1 |
| COVID19_63_R | GTGACTGGAGTTCAGACGTGTGCTCTTCCGATCTACAAACTGCCACCATCACAACC | Pool 1 |
| COVID19_64_F | TCTTTCCCTACACGACGCTCTTCCGATCTTCGATAGATATCCTGCTAATTCCATTGT | Pool 2 |
| COVID19_64_R | GTGACTGGAGTTCAGACGTGTGCTCTTCCGATCTAGTCTTGTAAAAGTGTTCCAGAGGT | Pool 2 |
| COVID19_65_F | TCTTTCCCTACACGACGCTCTTCCGATCTGCTGGCTTTAGCTTGTGGGTTT | Pool 1 |
| COVID19_65_R | GTGACTGGAGTTCAGACGTGTGCTCTTCCGATCTTGTCAGTCATAGAACAAACACCAATAGT | Pool 1 |
| COVID19_66_F | TCTTTCCCTACACGACGCTCTTCCGATCTGGGTGTGGACATTGCTGCTAAT | Pool 2 |
| COVID19_66_R | GTGACTGGAGTTCAGACGTGTGCTCTTCCGATCTTCAATTTCCATTTGACTCCTGGGT | Pool 2 |
| COVID19_67_F | TCTTTCCCTACACGACGCTCTTCCGATCTGTTGTCCAACAATTACCTGAAACTTACT | Pool 3 |
| COVID19_67_R | GTGACTGGAGTTCAGACGTGTGCTCTTCCGATCTCAACCTTAGAAACTACAGATAAATCTTGGG | Pool 3 |
| COVID19_68_F | TCTTTCCCTACACGACGCTCTTCCGATCTACAGGTTCATCTAAGTGTGTGTGT | Pool 2 |
| COVID19_68_R | GTGACTGGAGTTCAGACGTGTGCTCTTCCGATCTCTCCTTTATCAGAACCAGCACCA | Pool 2 |
| COVID19_69_F | TCTTTCCCTACACGACGCTCTTCCGATCTTGTCGCAAAATATACTCAACTGTGTCA | Pool 1 |
| COVID19_69_R | GTGACTGGAGTTCAGACGTGTGCTCTTCCGATCTTCTTTATAGCCACGGAACCTCCA | Pool 1 |

| | | |
|---|---|---|
| COVID19_70_F | TCTTTCCCTACACGACGCTCTTCCGATCTACAAAAGAAAATGACTCTAAAGAGGGTTT | Pool 2 |
| COVID19_70_R | GTGACTGGAGTTCAGACGTGTGCTCTTCCGATCTTGACCTTCTTTTAAAGACATAACAGCAG | Pool 2 |
| COVID19_71_F | TCTTTCCCTACACGACGCTCTTCCGATCTACAAATCCAATTCAGTTGTCTTCCTATTC | Pool 1 |
| COVID19_71_R | GTGACTGGAGTTCAGACGTGTGCTCTTCCGATCTTGGAAAAGAAAGGTAAGAACAAGTCCT | Pool 1 |
| COVID19_72_F | TCTTTCCCTACACGACGCTCTTCCGATCTACACGTGGTGTTTATTACCCTGAC | Pool 2 |
| COVID19_72_R | GTGACTGGAGTTCAGACGTGTGCTCTTCCGATCTACTCTGAACTCACTTTCCATCCAAC | Pool 2 |
| COVID19_73_F | TCTTTCCCTACACGACGCTCTTCCGATCTCAATTTTGTAATGATCCATTTTTGGGTGT | Pool 1 |
| COVID19_73_R | GTGACTGGAGTTCAGACGTGTGCTCTTCCGATCTCACCAGCTGTCCAACCTGAAGA | Pool 1 |
| COVID19_74_F | TCTTTCCCTACACGACGCTCTTCCGATCTACATCACTAGGTTTCAAACTTTACTTGC | Pool 3 |
| COVID19_74_R | GTGACTGGAGTTCAGACGTGTGCTCTTCCGATCTGCAACACAGTTGCTGATTCTCTTC | Pool 3 |
| COVID19_75_F | TCTTTCCCTACACGACGCTCTTCCGATCTAGAGTCCAACCAACAGAATCTATTGT | Pool 3 |
| COVID19_75_R | GTGACTGGAGTTCAGACGTGTGCTCTTCCGATCTACCACCAACCTTAGAATCAAGATTGT | Pool 3 |
| COVID19_76_F | TCTTTCCCTACACGACGCTCTTCCGATCTAGGGCAAACTGGAAAGATTGCT | Pool 3 |
| COVID19_76_F_alt3 | TCTTTCCCTACACGACGCTCTTCCGATCTGGGCAAACTGGAAAGATTGCTGA | Pool 3 |
| COVID19_76_R | GTGACTGGAGTTCAGACGTGTGCTCTTCCGATCTACACCTGTGCCTGTTAAACCAT | Pool 3 |
| COVID19_76_R_alt0 | GTGACTGGAGTTCAGACGTGTGCTCTTCCGATCTACCTGTGCCTGTTAAACCATTGA | Pool 3 |
| COVID19_77_F | TCTTTCCCTACACGACGCTCTTCCGATCTCCAGCAACTGTTTGTGGACCTA | Pool 1 |
| COVID19_77_R | GTGACTGGAGTTCAGACGTGTGCTCTTCCGATCTCAGCCCCTATTAAACAGCCTGC | Pool 1 |
| COVID19_78_F | TCTTTCCCTACACGACGCTCTTCCGATCTCAACTTACTCCTACTTGGCGTGT | Pool 2 |
| COVID19_78_R | GTGACTGGAGTTCAGACGTGTGCTCTTCCGATCTTGTGTACAAAAACTGCCATATTGCA | Pool 2 |
| COVID19_79_F | TCTTTCCCTACACGACGCTCTTCCGATCTGTGGTGATTCAACTGAATGCAGC | Pool 1 |

| | | |
|---|---|---|
| COVID19_79_R | GTGACTGGAGTTCAGACGTGTGCTCTTCCGATCTCATTTCATCTGTGAGCAAAGGTGG | Pool 1 |
| COVID19_80_F | TCTTTCCCTACACGACGCTCTTCCGATCTTTGCCTTGGTGATATTGCTGCT | Pool 2 |
| COVID19_80_R | GTGACTGGAGTTCAGACGTGTGCTCTTCCGATCTTGGAGCTAAGTTGTTTAACAAGCG | Pool 2 |
| COVID19_81_F | TCTTTCCCTACACGACGCTCTTCCGATCTGCACTTGGAAAACTTCAAGATGTGG | Pool 1 |
| COVID19_81_R | GTGACTGGAGTTCAGACGTGTGCTCTTCCGATCTGTGAAGTTCTTTTCTTGTGCAGGG | Pool 1 |
| COVID19_82_F | TCTTTCCCTACACGACGCTCTTCCGATCTGGGCTATCATCTTATGTCCTTCCCT | Pool 2 |
| COVID19_82_R | GTGACTGGAGTTCAGACGTGTGCTCTTCCGATCTTGCCAGAGATGTCACCTAAATCAA | Pool 2 |
| COVID19_83_F | TCTTTCCCTACACGACGCTCTTCCGATCTTCCTTTGCAACCTGAATTAGACTCA | Pool 1 |
| COVID19_83_R | GTGACTGGAGTTCAGACGTGTGCTCTTCCGATCTTTTGACTCCTTTGAGCACTGGC | Pool 1 |
| COVID19_84_F | TCTTTCCCTACACGACGCTCTTCCGATCTTGCTGTAGTTGTCTCAAGGGCT | Pool 2 |
| COVID19_84_R | GTGACTGGAGTTCAGACGTGTGCTCTTCCGATCTAGGTGTGAGTAAACTGTTACAAACAAC | Pool 2 |
| COVID19_85_F | TCTTTCCCTACACGACGCTCTTCCGATCTACTAGCACTCTCCAAGGGTGTT | Pool 1 |
| COVID19_85_R | GTGACTGGAGTTCAGACGTGTGCTCTTCCGATCTACACAGTCTTTTACTCCAGATTCCC | Pool 1 |
| COVID19_86_F | TCTTTCCCTACACGACGCTCTTCCGATCTTCAGGTGATGGCACAACAAGTC | Pool 2 |
| COVID19_86_R | GTGACTGGAGTTCAGACGTGTGCTCTTCCGATCTACGAAAGCAAGAAAAAGAAGTACGC | Pool 2 |
| COVID19_87_F | TCTTTCCCTACACGACGCTCTTCCGATCTCGACTACTAGCGTGCCTTTGTA | Pool 1 |
| COVID19_87_R | GTGACTGGAGTTCAGACGTGTGCTCTTCCGATCTACTAGGTTCCATTGTTCAAGGAGC | Pool 1 |
| COVID19_88_F | TCTTTCCCTACACGACGCTCTTCCGATCTCCATGGCAGATTCCAACGGTAC | Pool 2 |
| COVID19_88_R | GTGACTGGAGTTCAGACGTGTGCTCTTCCGATCTTGGTCAGAATAGTGCCATGGAGT | Pool 2 |
| COVID19_89_F | TCTTTCCCTACACGACGCTCTTCCGATCTGTACGCGTTCCATGTGGTCATT | Pool 1 |
| COVID19_89_F_alt2 | TCTTTCCCTACACGACGCTCTTCCGATCTCGCGTTCCATGTGGTCATTCAA | Pool 1 |

| | | |
|---|---|---|
| COVID19_89_R | GTGACTGGAGTTCAGACGTGTGCTCTTCCGATCTACCTGAAAGTCAACGAGATGAAACA | Pool 1 |
| COVID19_89_R_alt4 | GTGACTGGAGTTCAGACGTGTGCTCTTCCGATCTACGAGATGAAACATCTGTTGTCACT | Pool 1 |
| COVID19_90_F | TCTTTCCCTACACGACGCTCTTCCGATCTACACAGACCATTCCAGTAGCAGT | Pool 2 |
| COVID19_90_R | GTGACTGGAGTTCAGACGTGTGCTCTTCCGATCTTGAAATGGTGAATTGCCCTCGT | Pool 2 |
| COVID19_91_F | TCTTTCCCTACACGACGCTCTTCCGATCTTCACTACCAAGAGTGTGTTAGAGGT | Pool 1 |
| COVID19_91_R | GTGACTGGAGTTCAGACGTGTGCTCTTCCGATCTTTCAAGTGAGAACCAAAAGATAATAAGCA | Pool 1 |
| COVID19_92_F | TCTTTCCCTACACGACGCTCTTCCGATCTTTTGTGCTTTTTAGCCTTTCTGCT | Pool 2 |
| COVID19_92_R | GTGACTGGAGTTCAGACGTGTGCTCTTCCGATCTAGGTTCCTGGCAATTAATTGTAAAAGG | Pool 2 |
| COVID19_93_F | TCTTTCCCTACACGACGCTCTTCCGATCTTGAGGCTGGTTCTAAATCACCCA | Pool 1 |
| COVID19_93_R | GTGACTGGAGTTCAGACGTGTGCTCTTCCGATCTAGGTCTTCCTTGCCATGTTGAG | Pool 1 |
| COVID19_94_F | TCTTTCCCTACACGACGCTCTTCCGATCTGGCCCCAAGGTTTACCCAATAA | Pool 2 |
| COVID19_94_R | GTGACTGGAGTTCAGACGTGTGCTCTTCCGATCTTTTGGCAATGTTGTTCCTTGAGG | Pool 2 |
| COVID19_95_F | TCTTTCCCTACACGACGCTCTTCCGATCTTGAGGGAGCCTTGAATACACCA | Pool 1 |
| COVID19_95_R | GTGACTGGAGTTCAGACGTGTGCTCTTCCGATCTCAGTACGTTTTTGCCGAGGCTT | Pool 1 |
| COVID19_96_F | TCTTTCCCTACACGACGCTCTTCCGATCTGCCAACAACAACAAGGCCAAAC | Pool 2 |
| COVID19_96_R | GTGACTGGAGTTCAGACGTGTGCTCTTCCGATCTTAGGCTCTGTTGGTGGGAATGT | Pool 2 |
| COVID19_97_F | TCTTTCCCTACACGACGCTCTTCCGATCTTGGATGACAAAGATCCAAATTTCAAAGA | Pool 1 |
| COVID19_97_R | GTGACTGGAGTTCAGACGTGTGCTCTTCCGATCTACACACTGATTAAAGATTGCTATGTGAG | Pool 1 |
| COVID19_98_F | TCTTTCCCTACACGACGCTCTTCCGATCTAACAATTGCAACAATCCATGAGCA | Pool 2 |
| COVID19_98_R | GTGACTGGAGTTCAGACGTGTGCTCTTCCGATCTTTCTCCTAAGAAGCTATTAAAATCACATGG | Pool 2 |

**Supplementary Table 4:** Genotype at potentially mutated sites L9P, G172R, E484K, N501Y, A570D, D614G, P681H, and T716I determined by SIRPH analysis (SNuPE) and whole viral genome sequencing (Seq.). In sequencing data regions with coverage less than 20 were excluded (NA) from the analysis.

| ID | SNuPE L9P | SNuPE G172R | SNuPE E484K | SNuPE N501Y | SNuPE A570D | SNuPE D614G | SNuPE P681H | SNuPE T716I | Seq. L9P | Seq. G172R | Seq. E484K | Seq. N501Y | Seq. A570D | Seq. D614G | Seq. P681H | Seq. T716I |
|---|---|---|---|---|---|---|---|---|---|---|---|---|---|---|---|---|
| 2 | T (WT) | G (WT) | G (WT) | A (WT) | C (WT) | G (Mut) | C (WT) | C (WT) | T (WT) | G (WT) | G (WT) | A (WT) | C (WT) | G (Mut) | C (WT) | C (WT) |
| 3 | T (WT) | G (WT) | G (WT) | A (WT) | C (WT) | G (Mut) | C (WT) | C (WT) | T (WT) | G (WT) | G (WT) | A (WT) | C (WT) | G (Mut) | C (WT) | C (WT) |
| 4 | T (WT) | G (WT) | G (WT) | A (WT) | C (WT) | G (Mut) | C (WT) | C (WT) | T (WT) | G (WT) | NA | NA | C (WT) | G (Mut) | C (WT) | C (WT) |
| 5 | T (WT) | G (WT) | G (WT) | A (WT) | C (WT) | G (Mut) | C (WT) | C (WT) | T (WT) | G (WT) | G (WT) | A (WT) | C (WT) | G (Mut) | C (WT) | C (WT) |
| 6 | T (WT) | C (Mut) | G (WT) | A (WT) | C (WT) | G (Mut) | C (WT) | C (WT) | T (WT) | C (Mut) | G (WT) | A (WT) | C (WT) | G (Mut) | C (WT) | C (WT) |
| 7 | T (WT) | G (WT) | G (WT) | A (WT) | C (WT) | G (Mut) | C (WT) | C (WT) | T (WT) | G (WT) | NA | NA | C (WT) | G (Mut) | C (WT) | C (WT) |
| 8 | T (WT) | C (Mut) | G (WT) | A (WT) | C (WT) | G (Mut) | C (WT) | C (WT) | T (WT) | C (Mut) | G (WT) | A (WT) | C (WT) | G (Mut) | C (WT) | C (WT) |
| 9 | T (WT) | G (WT) | G (WT) | A (WT) | C (WT) | G (Mut) | C (WT) | C (WT) | T (WT) | G (WT) | G (WT) | NA | C (WT) | G (Mut) | C (WT) | C (WT) |
| 10 | T (WT) | C (Mut) | G (WT) | A (WT) | C (WT) | G (Mut) | C (WT) | C (WT) | T (WT) | C (Mut) | G (WT) | A (WT) | C (WT) | G (Mut) | C (WT) | C (WT) |
| 11 | T (WT) | G (WT) | G (WT) | A (WT) | C (WT) | G (Mut) | C (WT) | C (WT) | T (WT) | G (WT) | G (WT) | A (WT) | C (WT) | G (Mut) | C (WT) | C (WT) |
| 12 | T (WT) | C (Mut) | G (WT) | A (WT) | C (WT) | G (Mut) | C (WT) | C (WT) | T (WT) | NA | G (WT) | A (WT) | C (WT) | G (Mut) | C (WT) | C (WT) |
| 13 | T (WT) | G (WT) | G (WT) | A (WT) | C (WT) | G (Mut) | C (WT) | C (WT) | T (WT) | G (WT) | G (WT) | A (WT) | C (WT) | G (Mut) | C (WT) | C (WT) |
| 14 | T (WT) | C (Mut) | G (WT) | A (WT) | C (WT) | G (Mut) | C (WT) | C (WT) | T (WT) | NA | G (WT) | A (WT) | C (WT) | G (Mut) | C (WT) | C (WT) |
| 15 | C (Mut) | G (WT) | G (WT) | A (WT) | C (WT) | G (Mut) | C (WT) | C (WT) | NA | G (WT) | G (WT) | A (WT) | C (WT) | G (Mut) | C (WT) | C (WT) |
| 16 | C (Mut) | G (WT) | G (WT) | A (WT) | C (WT) | G (Mut) | C (WT) | C (WT) | NA | G (WT) | G (WT) | A (WT) | C (WT) | G (Mut) | C (WT) | C (WT) |
| 17 | T (WT) | G (WT) | G (WT) | A (WT) | C (WT) | G (Mut) | C (WT) | C (WT) | T (WT) | G (WT) | G (WT) | A (WT) | C (WT) | G (Mut) | C (WT) | C (WT) |

| | | | | | | | | | | | | | | | | |
|---|---|---|---|---|---|---|---|---|---|---|---|---|---|---|---|---|
| 18 | T (WT) | G (WT) | G (WT) | A (WT) | C (WT) | G (Mut) | C (WT) | C (WT) | T (WT) | G (WT) | G (WT) | A (WT) | C (WT) | G (Mut) | C (WT) | C (WT) |
| 20 | T (WT) | C (Mut) | G (WT) | A (WT) | C (WT) | G (Mut) | C (WT) | C (WT) | T (WT) | C (Mut) | G (WT) | A (WT) | C (WT) | G (Mut) | C (WT) | C (WT) |
| 22 | T (WT) | G (WT) | G (WT) | A (WT) | C (WT) | G (Mut) | C (WT) | C (WT) | T (WT) | G (WT) | G (WT) | A (WT) | C (WT) | G (Mut) | C (WT) | C (WT) |
| 23 | T (WT) | G (WT) | G (WT) | A (WT) | C (WT) | G (Mut) | C (WT) | C (WT) | T (WT) | G (WT) | G (WT) | A (WT) | C (WT) | G (Mut) | C (WT) | C (WT) |
| 24 | T (WT) | G (WT) | G (WT) | A (WT) | C (WT) | G (Mut) | C (WT) | C (WT) | T (WT) | G (WT) | G (WT) | A (WT) | C (WT) | G (Mut) | C (WT) | C (WT) |
| 25 | T (WT) | G (WT) | G (WT) | A (WT) | C (WT) | G (Mut) | C (WT) | C (WT) | T (WT) | G (WT) | G (WT) | A (WT) | C (WT) | G (Mut) | C (WT) | C (WT) |
| 26 | T (WT) | C (Mut) | G (WT) | A (WT) | C (WT) | G (Mut) | C (WT) | C (WT) | T (WT) | C (Mut) | NA | NA | C (WT) | G (Mut) | C (WT) | C (WT) |
| 27 | T (WT) | G (WT) | G (WT) | A (WT) | C (WT) | G (Mut) | C (WT) | C (WT) | NA | G (WT) | NA | NA | C (WT) | G (Mut) | C (WT) | C (WT) |
| 28 | T (WT) | G (WT) | G (WT) | A (WT) | C (WT) | G (Mut) | C (WT) | C (WT) | T (WT) | G (WT) | NA | NA | C (WT) | G (Mut) | C (WT) | C (WT) |
| 29 | T (WT) | C (Mut) | G (WT) | A (WT) | C (WT) | G (Mut) | C (WT) | C (WT) | T (WT) | C (Mut) | G (WT) | A (WT) | C (WT) | G (Mut) | C (WT) | C (WT) |
| 30 | T (WT) | G (WT) | G (WT) | A (WT) | C (WT) | G (Mut) | C (WT) | C (WT) | T (WT) | G (WT) | G (WT) | A (WT) | C (WT) | G (Mut) | C (WT) | C (WT) |
| 31 | T (WT) | G (WT) | G (WT) | A (WT) | C (WT) | G (Mut) | C (WT) | C (WT) | T (WT) | G (WT) | G (WT) | A (WT) | C (WT) | G (Mut) | C (WT) | C (WT) |
| 32 | C (Mut) | G (WT) | G (WT) | A (WT) | C (WT) | G (Mut) | C (WT) | C (WT) | C (Mut) | G (WT) | G (WT) | A (WT) | C (WT) | G (Mut) | C (WT) | C (WT) |
| 33 | T (WT) | G (WT) | G (WT) | A (WT) | C (WT) | G (Mut) | C (WT) | C (WT) | NA | G (WT) | G (WT) | A (WT) | C (WT) | G (Mut) | C (WT) | C (WT) |
| 34 | T (WT) | C (Mut) | G (WT) | A (WT) | C (WT) | G (Mut) | C (WT) | C (WT) | NA | NA | G (WT) | A (WT) | C (WT) | G (Mut) | C (WT) | C (WT) |
| 35 | T (WT) | C (Mut) | G (WT) | A (WT) | C (WT) | G (Mut) | C (WT) | C (WT) | T (WT) | NA | G (WT) | A (WT) | C (WT) | G (Mut) | C (WT) | C (WT) |
| 36 | T (WT) | C (Mut) | G (WT) | A (WT) | C (WT) | G (Mut) | C (WT) | C (WT) | NA | C (Mut) | G (WT) | A (WT) | C (WT) | G (Mut) | C (WT) | C (WT) |
| 37 | T (WT) | C (Mut) | G (WT) | A (WT) | C (WT) | G (Mut) | C (WT) | C (WT) | NA | NA | G (WT) | A (WT) | C (WT) | G (Mut) | C (WT) | C (WT) |
| 38 | T (WT) | C (Mut) | G (WT) | A (WT) | C (WT) | G (Mut) | C (WT) | C (WT) | NA | C (Mut) | G (WT) | A (WT) | C (WT) | G (Mut) | C (WT) | C (WT) |
| 39 | T (WT) | G (WT) | G (WT) | A (WT) | C (WT) | G (Mut) | C (WT) | C (WT) | NA | G (WT) | G (WT) | A (WT) | C (WT) | G (Mut) | C (WT) | C (WT) |
| 40 | T (WT) | G (WT) | G (WT) | A (WT) | C (WT) | G (Mut) | C (WT) | C (WT) | NA | G (WT) | G (WT) | A (WT) | C (WT) | G (Mut) | C (WT) | C (WT) |

| | | | | | | | | | | | | | | | | |
|---|---|---|---|---|---|---|---|---|---|---|---|---|---|---|---|---|
| 41 | T (WT) | G (WT) | G (WT) | A (WT) | C (WT) | G (Mut) | C (WT) | C (WT) | NA | G (WT) | G (WT) | A (WT) | C (WT) | G (Mut) | C (WT) | C (WT) |
| 42 | T (WT) | G (WT) | G (WT) | A (WT) | C (WT) | G (Mut) | C (WT) | C (WT) | NA | G (WT) | G (WT) | A (WT) | C (WT) | G (Mut) | C (WT) | C (WT) |
| 43 | T (WT) | G (WT) | G (WT) | A (WT) | C (WT) | G (Mut) | C (WT) | C (WT) | NA | G (WT) | G (WT) | A (WT) | C (WT) | G (Mut) | C (WT) | C (WT) |
| 44 | T (WT) | G (WT) | G (WT) | A (WT) | C (WT) | G (Mut) | C (WT) | C (WT) | NA | G (WT) | G (WT) | A (WT) | C (WT) | G (Mut) | C (WT) | C (WT) |
| 45 | T (WT) | G (WT) | G (WT) | A (WT) | C (WT) | G (Mut) | C (WT) | C (WT) | NA | G (WT) | G (WT) | A (WT) | C (WT) | G (Mut) | C (WT) | C (WT) |
| 46 | T (WT) | G (WT) | G (WT) | A (WT) | C (WT) | G (Mut) | C (WT) | C (WT) | NA | G (WT) | G (WT) | A (WT) | C (WT) | G (Mut) | C (WT) | C (WT) |
| 47 | T (WT) | G (WT) | G (WT) | A (WT) | C (WT) | G (Mut) | C (WT) | C (WT) | T (WT) | G (WT) | G (WT) | A (WT) | C (WT) | G (Mut) | C (WT) | C (WT) |
| 48 | T (WT) | G (WT) | G (WT) | A (WT) | C (WT) | G (Mut) | C (WT) | C (WT) | NA | G (WT) | G (WT) | A (WT) | C (WT) | G (Mut) | C (WT) | C (WT) |
| 49 | T (WT) | G (WT) | G (WT) | A (WT) | C (WT) | G (Mut) | C (WT) | C (WT) | NA | G (WT) | G (WT) | A (WT) | C (WT) | G (Mut) | C (WT) | C (WT) |
| 50 | T (WT) | G (WT) | G (WT) | A (WT) | C (WT) | G (Mut) | C (WT) | C (WT) | NA | G (WT) | G (WT) | A (WT) | C (WT) | G (Mut) | C (WT) | C (WT) |
| 51 | T (WT) | G (WT) | G (WT) | A (WT) | C (WT) | G (Mut) | C (WT) | C (WT) | T (WT) | G (WT) | G (WT) | A (WT) | C (WT) | G (Mut) | C (WT) | C (WT) |
| 53 | T (WT) | G (WT) | G (WT) | A (WT) | C (WT) | G (Mut) | C (WT) | C (WT) | T (WT) | G (WT) | G (WT) | A (WT) | C (WT) | G (Mut) | C (WT) | C (WT) |
| 54 | T (WT) | G (WT) | G (WT) | A (WT) | C (WT) | G (Mut) | C (WT) | C (WT) | NA | G (WT) | G (WT) | NA | C (WT) | G (Mut) | C (WT) | C (WT) |
| 55 | T (WT) | G (WT) | G (WT) | A (WT) | C (WT) | G (Mut) | C (WT) | C (WT) | NA | G (WT) | G (WT) | A (WT) | C (WT) | G (Mut) | C (WT) | C (WT) |
| 56 | T (WT) | G (WT) | G (WT) | A (WT) | C (WT) | G (Mut) | C (WT) | C (WT) | NA | G (WT) | G (WT) | A (WT) | C (WT) | G (Mut) | C (WT) | C (WT) |
| 57 | T (WT) | G (WT) | G (WT) | A (WT) | C (WT) | G (Mut) | C (WT) | C (WT) | T (WT) | G (WT) | NA | NA | C (WT) | G (Mut) | C (WT) | C (WT) |
| 58 | T (WT) | G (WT) | G (WT) | A (WT) | C (WT) | G (Mut) | C (WT) | C (WT) | T (WT) | G (WT) | NA | NA | C (WT) | G (Mut) | C (WT) | C (WT) |
| 59 | T (WT) | G (WT) | G (WT) | A (WT) | C (WT) | G (Mut) | C (WT) | C (WT) | NA | G (WT) | G (WT) | A (WT) | C (WT) | G (Mut) | C (WT) | C (WT) |
| 60 | T (WT) | C (Mut) | G (WT) | A (WT) | C (WT) | G (Mut) | C (WT) | C (WT) | NA | C (Mut) | G (WT) | A (WT) | C (WT) | G (Mut) | C (WT) | C (WT) |
| 61 | T (WT) | C (Mut) | G (WT) | A (WT) | C (WT) | G (Mut) | C (WT) | C (WT) | NA | C (Mut) | G (WT) | A (WT) | C (WT) | G (Mut) | C (WT) | C (WT) |
| 62 | T (WT) | C (Mut) | G (WT) | A (WT) | C (WT) | G (Mut) | C (WT) | C (WT) | NA | C (Mut) | G (WT) | A (WT) | C (WT) | G (Mut) | C (WT) | C (WT) |

| | | | | | | | | | | | | | | | | |
|---|---|---|---|---|---|---|---|---|---|---|---|---|---|---|---|---|
| 63 | T (WT) | C (Mut) | G (WT) | A (WT) | C (WT) | G (Mut) | C (WT) | C (WT) | NA | NA | G (WT) | A (WT) | C (WT) | G (Mut) | C (WT) | C (WT) |
| 64 | T (WT) | C (Mut) | G (WT) | A (WT) | C (WT) | G (Mut) | C (WT) | C (WT) | NA | C (Mut) | NA | NA | C (WT) | G (Mut) | C (WT) | C (WT) |
| 65 | T (WT) | G (WT) | G (WT) | A (WT) | C (WT) | G (Mut) | C (WT) | C (WT) | NA | G (WT) | NA | NA | C (WT) | G (Mut) | C (WT) | C (WT) |
| 66 | T (WT) | C (Mut) | G (WT) | A (WT) | C (WT) | G (Mut) | C (WT) | C (WT) | NA | C (Mut) | NA | NA | C (WT) | G (Mut) | C (WT) | C (WT) |
| 67 | T (WT) | C (Mut) | G (WT) | A (WT) | C (WT) | G (Mut) | C (WT) | C (WT) | T (WT) | C (Mut) | NA | NA | C (WT) | G (Mut) | C (WT) | C (WT) |
| 68 | T (WT) | G (WT) | G (WT) | A (WT) | C (WT) | G (Mut) | C (WT) | C (WT) | T (WT) | G (WT) | G (WT) | NA | C (WT) | G (Mut) | C (WT) | C (WT) |
| 69 | T (WT) | C (Mut) | G (WT) | A (WT) | C (WT) | G (Mut) | C (WT) | C (WT) | NA | C (Mut) | G (WT) | A (WT) | C (WT) | G (Mut) | C (WT) | C (WT) |
| 70 | T (WT) | G (WT) | G (WT) | A (WT) | C (WT) | G (Mut) | C (WT) | C (WT) | T (WT) | G (WT) | G (WT) | A (WT) | C (WT) | G (Mut) | C (WT) | C (WT) |
| 71 | T (WT) | G (WT) | G (WT) | A (WT) | C (WT) | G (Mut) | C (WT) | C (WT) | T (WT) | G (WT) | G (WT) | A (WT) | C (WT) | G (Mut) | C (WT) | C (WT) |
| 72 | T (WT) | G (WT) | G (WT) | A (WT) | C (WT) | G (Mut) | C (WT) | C (WT) | NA | G (WT) | G (WT) | A (WT) | C (WT) | G (Mut) | C (WT) | C (WT) |
| 73 | T (WT) | C (Mut) | G (WT) | A (WT) | C (WT) | G (Mut) | C (WT) | C (WT) | T (WT) | C (Mut) | G (WT) | A (WT) | C (WT) | G (Mut) | C (WT) | C (WT) |
| 74 | T (WT) | G (WT) | G (WT) | A (WT) | C (WT) | G (Mut) | C (WT) | C (WT) | NA | G (WT) | G (WT) | A (WT) | C (WT) | G (Mut) | C (WT) | C (WT) |
| 75 | T (WT) | G (WT) | G (WT) | A (WT) | C (WT) | G (Mut) | C (WT) | C (WT) | NA | G (WT) | G (WT) | A (WT) | C (WT) | G (Mut) | C (WT) | C (WT) |
| 76 | T (WT) | G (WT) | G (WT) | A (WT) | C (WT) | G (Mut) | C (WT) | C (WT) | NA | G (WT) | G (WT) | A (WT) | C (WT) | G (Mut) | C (WT) | C (WT) |
| 77 | T (WT) | G (WT) | G (WT) | A (WT) | C (WT) | G (Mut) | C (WT) | C (WT) | T (WT) | G (WT) | NA | NA | C (WT) | G (Mut) | C (WT) | C (WT) |
| 78 | T (WT) | G (WT) | G (WT) | A (WT) | C (WT) | G (Mut) | C (WT) | C (WT) | NA | G (WT) | G (WT) | A (WT) | C (WT) | G (Mut) | C (WT) | C (WT) |
| 102 | T (WT) | G (WT) | G (WT) | A (WT) | C (WT) | G (Mut) | C (WT) | C (WT) | NA | G (WT) | NA | NA | C (WT) | G (Mut) | C (WT) | C (WT) |
| 108 | C (Mut) | G (WT) | G (WT) | A (WT) | C (WT) | G (Mut) | C (WT) | C (WT) | NA | G (WT) | NA | NA | C (WT) | G (Mut) | C (WT) | C (WT) |
| 109 | C (Mut) | G (WT) | G (WT) | A (WT) | C (WT) | G (Mut) | C (WT) | C (WT) | C (Mut) | G (WT) | G (WT) | A (WT) | C (WT) | G (Mut) | C (WT) | C (WT) |
| 110 | T (WT) | G (WT) | G (WT) | A (WT) | C (WT) | G (Mut) | C (WT) | C (WT) | NA | G (WT) | NA | NA | C (WT) | G (Mut) | C (WT) | C (WT) |
| 111 | T (WT) | G (WT) | G (WT) | A (WT) | C (WT) | G (Mut) | C (WT) | C (WT) | T (WT) | G (WT) | G (WT) | A (WT) | C (WT) | G (Mut) | C (WT) | C (WT) |

| | | | | | | | | | | | | | | | | |
|---|---|---|---|---|---|---|---|---|---|---|---|---|---|---|---|---|
| 112 | T (WT) | G (WT) | G (WT) | A (WT) | C (WT) | G (Mut) | C (WT) | C (WT) | T (WT) | G (WT) | NA | NA | C (WT) | G (Mut) | C (WT) | C (WT) |

**Supplementary Table 5:** Mutation profile of VOC candidates determined by SIRPH analysis.

| ID | SARS-CoV-2 PCR Ct | SNuPE (E484K) | SNuPE (N501Y) | SNuPE (A570D) | SNuPE (D614G) | SNuPE (P681H) | SNuPE (T716I) | SNuPE (V1176F) | VOC |
|---|---|---|---|---|---|---|---|---|---|
| 3004631047 | 15 | G (WT) | T (Mut) | A (Mut) | G (Mut) | A (Mut) | T (Mut) | NA | B1.1.7 |
| 3004631647 | 18 | G (WT) | T (Mut) | A (Mut) | G (Mut) | A (Mut) | T (Mut) | NA | B1.1.7 |
| 3004627147 | 19 | G (WT) | T (Mut) | A (Mut) | G (Mut) | A (Mut) | T (Mut) | NA | B1.1.7 |
| 3004631847 | 23 | G (WT) | T (Mut) | A (Mut) | G (Mut) | A (Mut) | T (Mut) | NA | B1.1.7 |
| 20291038 | 24 | G (WT) | T (Mut) | A (Mut) | G (Mut) | A (Mut) | T (Mut) | NA | B1.1.7 |
| 30046507 | 25 | G (WT) | T (Mut) | A (Mut) | G (Mut) | A (Mut) | T (Mut) | NA | B1.1.7 |
| 21003884 | 27 | G (WT) | T (Mut) | A (Mut) | G (Mut) | A (Mut) | T (Mut) | NA | B1.1.7 |
| 21003581 | 27 | G (WT) | T (Mut) | A (Mut) | G (Mut) | A (Mut) | T (Mut) | NA | B1.1.7 |
| 21003582 | 28 | G (WT) | T (Mut) | A (Mut) | G (Mut) | A (Mut) | T (Mut) | NA | B1.1.7 |
| 10474388 | 31 | A (Mut) | NA | C (WT) | G (Mut) | C (WT) | C (WT) | G (WT) | B.1.351 |
| 10474400 | NA | G (WT) | T (Mut) | A (Mut) | G (Mut) | A (Mut) | T (Mut) | NA | B1.1.7 |
| 10474401 | NA | G (WT) | T (Mut) | A (Mut) | G (Mut) | A (Mut) | T (Mut) | NA | B1.1.7 |

**Supplementary Table 6:** GISAID Identifiers for viral genome sequences. Note that only sequences with high quality (more than 95% genomic coverage with at least 20X) were submitted to GISAID. Consensus sequences with more missing sites but sufficient coverage at the analyzed potentially mutated sites can be found in Sup. Data 1.

| | | | | |
|---|---|---|---|---|
| EPI_ISL_1091121 | EPI_ISL_1091106 | EPI_ISL_1091114 | EPI_ISL_1091138 | EPI_ISL_1091093 |
| EPI_ISL_1091111 | EPI_ISL_1091120 | EPI_ISL_1091130 | EPI_ISL_1091103 | EPI_ISL_1091129 |
| EPI_ISL_1091122 | EPI_ISL_1091091 | EPI_ISL_1091126 | EPI_ISL_1091109 | EPI_ISL_1091135 |
| EPI_ISL_1091097 | EPI_ISL_1091101 | EPI_ISL_1091118 | EPI_ISL_1091136 | EPI_ISL_1091137 |
| EPI_ISL_1091115 | EPI_ISL_1091127 | EPI_ISL_1091125 | EPI_ISL_1091092 | EPI_ISL_1091100 |
| EPI_ISL_1091146 | EPI_ISL_1091108 | EPI_ISL_1091107 | EPI_ISL_1091098 | EPI_ISL_1091102 |
| EPI_ISL_1091099 | EPI_ISL_1091112 | EPI_ISL_1091140 | EPI_ISL_1091104 | EPI_ISL_1091094 |
| EPI_ISL_1091090 | EPI_ISL_1091096 | EPI_ISL_1091113 | EPI_ISL_1091117 | |
